# Effects of COVID-19 home confinement on physical activity and eating behaviour Preliminary results of the ECLB-COVID19 international online-survey

**DOI:** 10.1101/2020.05.04.20072447

**Authors:** Achraf Ammar, Michael Brach, Khaled Trabelsi, Hamdi Chtourou, Omar Boukhris, Liwa Masmoudi, Bassem Bouaziz, Ellen Bentlage, Daniella How, Mona Ahmed, Patrick Mueller, Notger Mueller, Asma Aloui, Omar Hammouda, Laisa Liane Paineiras-Domingos, Annemarie Braakman-jansen, Christian Wrede, Sophia Bastoni, Carlos Soares Pernambuco, Leonardo Mataruna, Morteza Taheri, Khadijeh Irandoust, Aïmen Khacharem, Nicola L Bragazzi, Karim Chamari, Jordan M Glenn, Nicholas T Bott, Faiez Gargouri, Lotfi Chaari, Hadj Batatia, Gamal Mohamed Ali, Osama Abdelkarim, Mohamed Jarraya, Kais El Abed, Nizar Souissi, Lisette Van Gemert-Pijnen, Bryan L Riemann, Laurel Riemann, Wassim Moalla, Jonathan Gómez-Raja, Monique Epstein, Robbert Sanderman, Sebastian Schulz, Achim Jerg, Ramzi Al-Horani, Taysir Mansi, Mohamed Jmail, Fernando Barbosa, Fernando Santos, Boštjan Šimunič, Rado Pišot, Donald Cowan, Andrea Gaggioli, Stephen J Bailey, Jürgen Steinacker, Tarak Driss, Anita Hoekelmann

**Affiliations:** Institute of Sport Science, Otto-von-Guericke University, 39106, Magdeburg, Germany; Institute of Sport and Exercise Sciences, Münster, Germany Michael Brach; High Institute of Sport and Physical Education of Sfax, 3000, Sfax, Tunisia; Higher Institute of Computer Science and Multimedia of Sfax, 3000, Sfax, Tunisia; Research Group Neuroprotection, German Center for Neurodegenerative Diseases (DZNE), Leipziger Str. 44, Magdeburg, 39120, Germany; Laboratório de Vibrações Mecânicas e Práticas Integrativas, Departamento de Biofísica e Biometria, Instituto de Biologia Roberto Alcântara Gomes e Policlínica Piquet Carneiro, Universidade do Estado do Rio de Janeiro, Rio de Janeiro, RJ, Brazil; University of Twente, the Netherlands Région de Enschede, Netherland; Catholic University of the Sacred Heart I UNICATT, Milano, Italy; Laboratório de Biociências da Motricidade Humana (LABIMH) da Universidade Federal do Estado do Rio de Janeiro (UNIRIO) – Rio de Janeiro/RJ – Brasil; College of Business Administration, American University in the Emirates, Dubai, UAE; Imam Khomeini International University, Qazvin, Iran; UVHC, DeVisu, Valenciennes, France; LIRTES - EA 7313. Université Paris Est Créteil Val de Marne; Department of Health Sciences (DISSAL), Postgraduate School of Public Health, University of Genoa, Genoa 16132, Italy; Department of Research and Education / Aspetar, Qatar; Exercise Science Research Center, Department of Health, Human Performance and Recreation, University of Arkansas, AR 72701, Fayetteville, USA; Clinical Excellence Research Center, Department of Medicine, Stanford University School of Medicine, CA 94305, Stanford, USA; University of Toulouse, IRIT - INP-ENSEEIHT, France; Faculty of Physical Education, Assiut University, Assiut 71515, Egypt; Karlsruher Institut für Technologie, Karlsruher, Germany; Activité Physique, Sport et Santé, UR18JS01, Observatoire National du Sport, Tunis, Tunisie; Georgia Southern University, Statesboro, GA 30458, USA; PharmD, BCBS; PharmIAD, Inc, Savannah, GA, USA; Health and Social Services, Fundesalud, 06800, Merida, Spain; The E-senior association, 75020 Paris, France; Department of Health Psychology, University Medical Center Groningen,University of Groningen, Groningen, The Netherlands; Department of Medicine, Ulm University, Leimgrubenweg 14, 89075 Ulm, Germany; Department of Exercise Science, Yarmouk University, Irbid, Jordan; Department of Instruction and Supervision, The University of Jordan, Jordan; Digital Research Centre of Sfax, Sfax, Tunisia; Faculty of Psychology and Education Sciences, University of Porto, Porto Portugal; ISCTE-Instituto Universitário de Lisboa, Av. das Forças Armadas, 1649-026 Lisboa, Portugal; Institute for Kinesiology Research, Science and Research Centre, Koper, Slovenia; Centre for Bioengineering and Biotechnology University of Waterloo, Waterloo, Canda; School of Sport, Exercise and Health Sciences, Loughborough University, Loughborough, UK; Interdisciplinary Laboratory in Neurosciences, Physiology and Psychology: Physical Activity, Health and Learning (LINP2-2APS), UFR STAPS, UPL, Paris Nanterre University, 92000 Nanterre, France

**Keywords:** Pandemic, Public health, Physical activity, Nutrition, COVID-19

## Abstract

**Background:** Public health recommendations and governmental measures during the COVID-19 pandemic have enforced numerous restrictions on daily living including social distancing, isolation and home confinement. While these measures are imperative to abate the spreading of COVID-19, the impact of these restrictions on health behaviours and lifestyle at home is undefined. Therefore, an international online survey was launched in April 2020 in seven languages to elucidate the behavioral and lifestyle consequences of COVID-19 restrictions. This report presents the preliminary results from the first thousand responders on physical activity (PA) and nutrition behaviours.

**Methods:** Thirty-five research organisations from Europe, North-Africa, Western Asia and the Americas promoted the survey through their networks to the general society, in English, German, French, Arabic, Spanish, Portugese, and Slovenian languages. Questions were presented in a differential format with questions related to responses “before” and “during” confinement conditions.

**Results:** 1047 replies (54% women) from Asia (36%), Africa (40%), Europe (21%) and other (3%) were included into a general analysis. The COVID-19 home confinement had a negative effect on all intensities of PA (vigorous, moderate, walking and overall). Conversely, daily sitting time increased from 5 to 8 hours per day. Additionally, food consumption and meal patterns (the type of food, eating out of control, snacks between meals, number of meals) were more unhealthy during confinement with only alcohol binge drink decreasing significantly.

**Conclusion:** While isolation is a necessary measure to protect public health, our results indicate that it alters physical activity and eating behaviours in a direction that would compromise health. A more detailed analysis of survey data will allow for a segregation of these responses in different age groups, countries and other subgroups which will help develop bespoke interventions to mitigate the negative lifestyle behaviors manifest during the COVID-19 confinement.

## Background

In the face of the present COVID-19 pandemic, public health recommendations and governmental measures have enforced lock-downs and restrictions. While these restrictions help to abate the rate of infection, such limitations result in negative effects by limiting participation in normal daily activities, physical activity (PA), travel and access to many forms of exercise (e.g., closed gyms, no groups gatherings, increased social distancing).^1^ Several countries are enforcing curfews, which limit the time to participate in outdoor activities, or are excluding outdoor activities entirely. Such restrictions impose a burden on the health in the population by potentially compromising physical fitness, which is positively associated with the ability to cope with an infection, and the immunologic and cardiopulmonary complications of more severe outcomes.^2,3^

Physical inactivity and poor mental health are among the most important risk factors for major disease morbidity, both in Western societies and globally.^4^ This holds true not only for the general population, but also for older adults and chronically ill patient populations who are at heightened risk of COVID-19-inducd ill health and mortality. For children and youth, physical activity is closely coupled to school-related activities, active transport and sport participation.^5^ Since schools have been closed during the COVID-19 pandemic this will also compromise physical activity in this population and might also risk longer-term sedentary behaviours.

In addition to the challenged to engage in PA, the closure of food suppliers has placed a burden on normal food-related behaviour.^6^ This is noteworthy since good nutrition is important for health and well-being, particularly when the immune system is challenged^7^ and limited access to fresh food could negatively affect overall physical and mental health.^8^ Anxiety and boredom evoked by quarantine are considered risk factors for consuming more food and food of a poorer quality compared to standard living conditions.^9^ Combined with the potential for lower levels of PA, consuming a high volume of poorer quality food would lead to a more positive energy balance.^10^

Currently, there is limited evidence to evaluate the effect of confinement on PA and diet behaviours. It is important to investigate how PA and eating behaviour can be affected by lengthy restrictions to establish a fundamental knowledge basis from which to inform appropriate recommendations, including lifestyle modifications during this time. Thus, the main research questions are:

1. To what extent is physical activity and eating behaviours changed during lengthy restrictions?
2. Are there changes in mental state, mood and sleep during lengthy restrictions?
3. Which demographic, cultural and lifestyle factors should be considered as moderators of these effects?

The present paper presents preliminary data on physical activity and nutrition responses before and during home confinement, collected by an international online survey (ECLB-COVID19, [link](https://docs.google.com/forms/d/e/1FAIpQLSfjapSdW80-Y8HLlDEreVbjHD_Za0ff_AjfGWhyWHns4dshOQ/viewform?fbzx=-9209079818555779482)). Other parts of the survey evaluate socialisation, sleep, mental and mood status, and general lifestyle and these findings will be published elsewhere. There is a common method description in all papers.

## Methods

We report findings on the first 1047 replies to an international online-survey on mental health and multi-dimension lifestyle behaviors during home confinement ([ECLB-COVID19](https://docs.google.com/forms/d/e/1FAIpQLSfjapSdW80-Y8HLlDEreVbjHD_Za0ff_AjfGWhyWHns4dshOQ/formResponse)). ECLB-COVID19 was opened on April 1, 2020, tested by the project’s steering group for a period of 1 week, before starting to spread it worldwide on April 6, 2020. Thirty-five research organizations from Europe, North-Africa, Western Asia and the Americas promoted dissemination and administration of the survey. ECLB-COVID19 was administered in English, German, French, Arabic, Spanish, Portuguese, and Slovenian languages (is currently provided also in Dutch, Persian and Italian). The survey included sixty-four questions on health, mental wellbeing, mood, life satisfaction and multidimension lifestyle behaviors (physical activity, diet, social participation, sleep, technology-use, need of psychosocial support). All questions were presented in a differential format, to be answered directly in sequence regarding “before” and “during” confinement conditions. The study was conducted according to the Declaration of Helsinki. The protocol and the consent form were fully approved (identification code: 62/20) by the Otto von Guericke University Ethics Committee.

## Survey development and promotion

The ECLB-COVID19 electronic survey was designed by a steering group of multidisciplinary scientists and academics (i.e., human science, sport science, neuropsychology and computer science) at the University of Magdeburg (principal investigator), the University of Sfax, the University of Münster and the University of Paris-Nanterre, following a structured review of the literature. The survey was then reviewed and edited by Over 50 colleagues and experts worldwide. The survey was uploaded and shared on the Google online survey platform. A link to the electronic survey was distributed worldwide by consortium colleagues via a range of methods: invitation via e-mails, shared in consortium’s faculties official pages, ResearchGate™, LinkedIn™ and other social media platforms such as Facebook™, WhatsApp™ and Twitter™. Public were also involved in the dissemination plans of our research through the promotion of the ECLB-COVID19 survey in their networks. The survey included an introductory page describing the background and the aims of the survey, the consortium, ethics information for participants and the option to choose one of seven available languages (English, German, French, Arabic, Spanish, Portuguese, and Slovenian). The present study focusses on the first thousand responses (i.e., 1047 participants), which were reached on April 11, 2020, approximately one-week after the survey began. This survey was open for all people worldwide aged 18 years or older. People with cognitive decline are excluded. Since the data were captured using Google Forms, participants were also required to read and confirm acceptance of Google’s privacy policy (https://policies.google.com/privacy?hl=en). Individuals were instructed that, by completing the survey, they were consenting to participate anonymously in the study.

## Survey questionnaires

The ECLB-COVID19 is a multi-country electronic survey designed to assess change in multiple lifestyle behaviors during the COVID-19 outbreak. Therefore, a collection of validated and/or crisis-oriented briefs questionnaires were included. These questionnaires assess mental wellbeing (Short Warwick-Edinburgh Mental Well-being Scale (SWEMWBS)),^11^ mood and feeling (Short Mood and Feelings Questionnaire (SMFQ)),^12^ life satisfaction (Short Life Satisfaction Questionnaire for Lockdowns (SLSQL), social participation (Short Social Participation Questionnaire for Lockdowns (SSPQL)), physical activity (International Physical Activity Questionnaire Short Form (IPAQ-SF)),^13,14^ diet behaviors (Short Diet Behaviors Questionnaire for Lockdowns (SDBQL)), sleep quality (Pittsburgh Sleep Quality Index (PSQI))^15^ and some key questions assessing the technology-use behaviors (Short Technology-use Behaviors Questionnaire for Lockdowns (STBQL)), demographic information and the need of psychosocial support. Reliability of the shortened and/or newly adopted questionnaires was tested by the project steering group through piloting, prior to survey administration. These brief crisis-oriented questionnaires showed good to excellent test-retest reliability coefficients (r = 0.84-0.96). A multi-language validated version already existed for the majority of these questionnaires and/or questions. However, for questionnaires that did not already exist in multi-language versions, we followed the procedure of translation and backtranslation, with an additional review for all language versions from the international scientists of our consortium. As a result, a total number of sixty-for items were included in the ECLB-COVID19 online survey in a differential format; that is, each item or question requested two answers, one regarding the period before and the other regarding the period during confinement. Thus, the participants were guided to compare the situations. Given the large number of questions included, the present paper focuses on the IPAQ-SF and the newly developed SDBQL as brief crisis-oriented tools.

### International Physical Activity Questionnaire Short Form (IPAQ-SF)

According to the official IPAQ-SF guidelines, data from the IPAQ-SF are summed within each item (i.e., vigorous intensity, moderate intensity, and walking) to estimate the total amount of time spent engaged in PA per week.^13,14^ Total weekly PA (MET-min week^-1^) was estimated by adding the products of reported time for each item by a MET value that was specific to each category of PA. We assigned 2 different sets of MET values. The first set was the original values (original IPAQ) based on the official IPAQ guidelines: vigorous PA = 8.0 METs, moderate PA = 4.0 METs, and walking = 3.3 METs. The other set used modified values (modified IPAQ), which we had devised for use with elderly adults, as reported by Stewart et al.^16^ and Yasunaga et al.^17^: vigorous PA = 5.3 METs, moderate PA = 3.0 METs, and walking = 2.5 METs. Additionally, we added the total PA (sum of performed vigorous, moderate and walking activity) as a 4^th^ item and sitting time as 5^th^ item in the present work.

### Short Diet Behavior Questionnaire for Lockdowns

The SDBQ-L is a newly developed crisis-oriented short questionnaire to assess dietary behavior before and during the lockdown period. The SDBQ-L has 5 questions related to “unhealthy food”, “eating out of control”, “snacks between meals”, “alcohol binge drinking”, and “number of meals/day” in parts referring to the Nutricalc questionnaire, Swiss society of nutrition (doi/10.1371/journal.pone.0143293.s003). The response choices and their designated scores were as follows: “Never” = 0; “Sometimes” = 1; “Most of the time” = 2; “Always” = 3. These choices and points were applied for the first four questionnaires. The choices and the designated scores for the fifth questionnaire were as follows: “1-2” = 1; “3” = 0; “4” = 1; “5” = 2; “>5” = 3. Total score of this questionnaire, corresponded to the sum of the scores in the 5 questions. The total score for the SDBQ-L is from “0” to “15”, where “0” designates no healthy dietary behaviors and “15” designates severe unhealthy dietary behaviours.

## Data Analysis

Descriptive statistics were used to define the proportion of responses for each question and the total distribution in the total score of each questionnaire. All statistical analyses were performed using the commercial statistical software STATISTICA (StatSoft, Paris, France, version 10.0) and Microsoft Excel 2010. Normality of the data distribution was confirmed using the Shapiro-Wilks-W-test. Values were computed and reported as mean ± SD (standard deviation). To assess for significant differences in responses “before” and “during” the confinement period, paired samples *t*-tests were used. Effect size (Cohen’s d) was calculated to determine the magnitude of the change of the score and was interpreted using the following criteria: 0.2 (small), 0.5 (moderate), and 0.8 (large).^18^ Statistical significance was accepted as p<0.05.

## Results

### Sample description

The present study focused on the first thousand responses (i.e., 1047 participants), which was reached on April, 11, 2020. Overall 54% of the participants were female women and the participants were from Asian (36%, mostly from Western Asia), African (40%, mostly from North Africa), European (21%) and other (3%) countries. Age, health status, employment status, level of education and marital status are presented in Table 1.

**Table 1:** Demographic characteristics of the participants.

| **Variables** | **N** | **(%)** |
| --- | --- | --- |
| **Gender** |  |  |
| Male | 484 | (46.2%) |
| Female | 563 | (53.8%) |
| **Continent** |  |  |
| North Africa | 419 | (40%) |
| Western Asia | 377 | (36%) |
| Europe | 220 | (21%) |
| Other | 31 | (3%) |
| **Age (years)** |  |  |
| 18-35 | 577 | (55.1%) |
| 36-55 | 367 | (35.1%) |
| >55 | 103 | (9.8%) |
| **Level of Education** |  |  |
| Master/doctorate degree | 527 | (50.3%) |
| Bachelor's degree | 397 | (37.9%) |
| Professional degree | 28 | (2.7%) |
| High school graduate, diploma or the equivalent | 69 | (6.6%) |
| No schooling completed | 26 | (2.5%) |
| **Marital status** |  |  |
| Single | 455 | (43.5%) |
| Married/Living as couple | 562 | (53.7%) |
| Widowed/Divorced/Separated | 30 | (2.9%) |
| **Employment status** |  |  |
| Employed for wages | 538 | (51.4%) |
| Self-employed | 74 | (7.1%) |
| Out of work/Unemployed | 75 | (7.2%) |
| A student | 259 | (24.7%) |
| Retired | 23 | (2.2%) |
| Unable to work | 9 | (0.9%) |
| Problem caused by COVID-19 | 59 | (5.6%) |
| Other | 10 | (1%) |
| **Health state** |  |  |
| Healthy | 956 | (91.3%) |
| With risk factors for cardiovascular disease | 81 | (7.7%) |
| With cardiovascular disease | 10 | (1%) |

### Physical activity before and during the confinement period

Responses to the physical activity questionnaire recorded before and during home confinement are presented in Table 2.

**Table 2.** Responses to the physical activity questionnaire recorded before and during home confinement.

| | | Before<br>confinement | During<br>confinement | $\Delta$ ($\Delta\%$) | t test | p value | Cohen's d |
| --- | --- | --- | --- | --- | --- | --- | --- |
| Vigorous<br>intensity | Days/week | 1.97±2.11 | 1.52±2.03 | 0.45 (22.7%) | 7.751 | <.001 | 0.374 |
| | min/week | 38.7±58.1 | 26.0±47.8 | 12.83 (33.1%) | 9.759 | <.001 | 0.542 |
| | MET values | 1168±2468.7 | 737.2±1844.5 | 430.8 (36.9%) | 6.683 | <.001 | 0.315 |
| Moderate<br>intensity | Days/week | 1.79±2.08 | 1.36±1.95 | 0.43 (24.0%) | 7.898 | <.001 | 0.396 |
| | min/week | 32.1±49 | 21.4±37.3 | 10.72 (33.4%) | 7.858 | <.001 | 0.343 |
| | MET values | 446.4±920.2 | 291.5±772.7 | 154.8 (34.7%) | 5.247 | <.001 | 0.204 |
| Walking | Days/week | 3.59±2.58 | 2.33±2.48 | 1.26 (35.0%) | 15.806 | <.001 | 0.677 |
| | min/week | 37.2±46.8 | 24.6±34.1 | 12.64 (34.0%) | 9.343 | <.001 | 0.389 |
| | MET values | 578.3±917.1 | 331.4±640.2 | 246.8 (42.7%) | 9.039 | <.001 | 0.361 |
| All PA | Days/week | 5.04±2.51 | 3.83±2.82 | 1.21 (24.0%) | 15.611 | <.001 | 0.482 |
| | min/week | 108±114.2 | 71.8±88.2 | 36.19 (33.5%) | 12.510 | <.001 | 0.387 |
| | MET values | 2192.6±3300.7 | 1360.2±2545.2 | 832.5 (38.0%) | 9.146 | <.001 | 0.283 |
| Sitting | Hours/day | 5.31±3.65 | 8.41±5.11 | 3.10 (28.6%) | -25.030 | <0.001 | 1.130 |

#### Vigorous intensity

The number of days/week and minutes/day of vigorous intensity PA during, compared to before, home confinement decreased by 22.7% (t=7.75, p<0.001, d=0.374) and 33.1% (t=9.75p<0.001, d=0.542). Additionally, MET values of vigorous intensity PA were 36.9% lower during, compared to before. home confinement (t=6.68, p<0.001, d=0.315).

#### Moderate intensity

The number of days/week of moderate intensity PA decreased by 24% during home confinement (t=7.89, p<0.001, d=0.396). Likewise, the number of minutes/day of moderate intensity decreased by 33.4% during home confinement (t=7.85, p<0.001, d=0.343). Additionally, MET values of moderate intensity were 34.7% lower during home confinement (t=5.24, p<0.001, d=0.204).

#### Walking

The number of days/week of walking decreased by 35% during home confinement (t=15.80, p<0.001, d=0.677). Likewise, the number of minutes/day of walking decreased by 34% during home confinement (t=9.34, p<0.001, d=0.389). Additionally, MET values of walking were 42.7% lower during home confinement (t=9.03, p<0.001, d=0.361).

#### All physical activity (PA)

The number of days/week of all PA decreased by 24% during home confinement (t=15.61, p<0.001, d=0.482). Likewise, the number of minutes/day of all PA decreased by 33.5% during home confinement (t=12.51, p<0.001, d=0.387). Additionally, MET values of all PA were 38% lower during home confinement (t=9.14, p<0.001, d=0.283).

#### Sitting

Statistical analysis reported that the number of hours/day of sitting increased by 28.6% during home confinement (t=-25.61, p<0.001, d=1.130).

### Dietary behaviors before and during the confinement period

Recorded scores in responses to the diet behavior questionnaire before and during home confinement are presented in Figure 1 (question 1 to 4) and Figure 2 (question 5).

**Figure 1:**
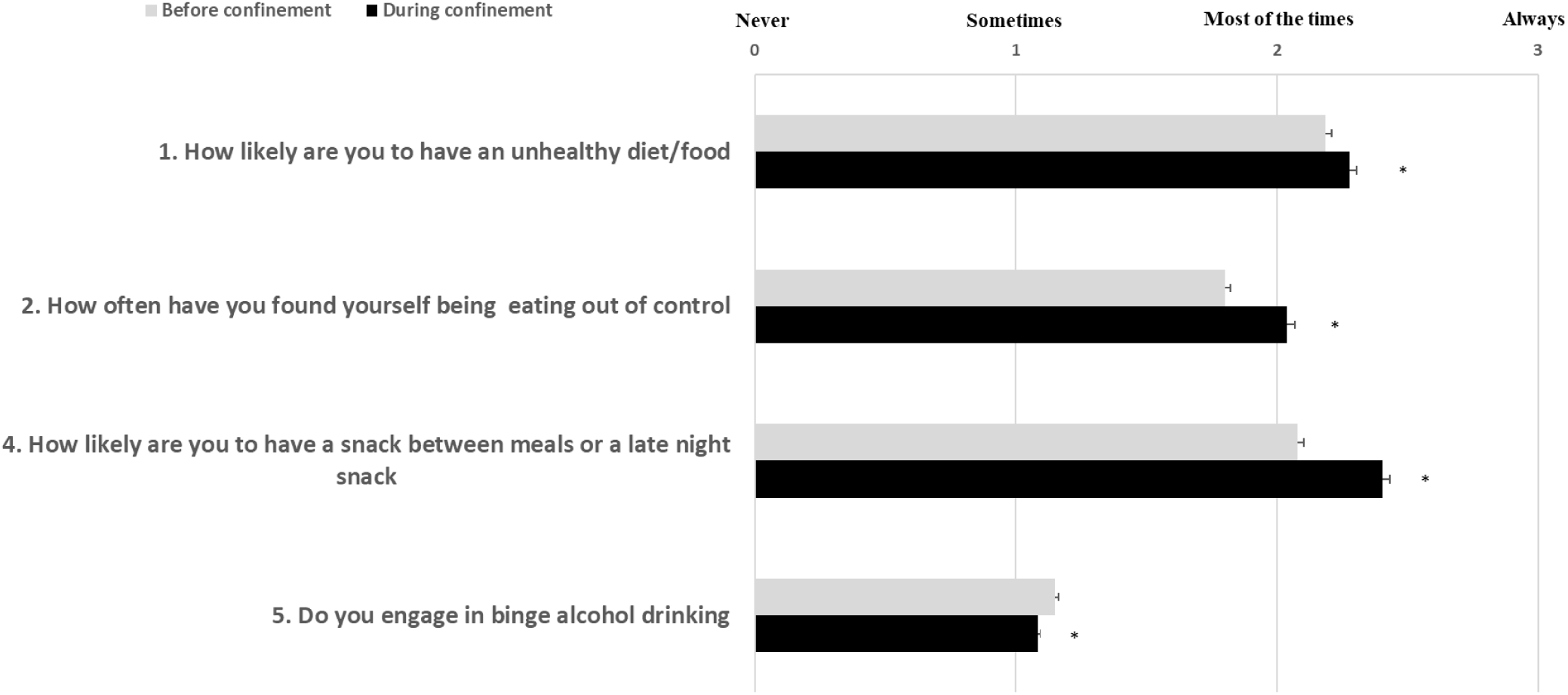
Participants’ scores in response to the related-diet behaviors’ questions.

**Figure 2:**
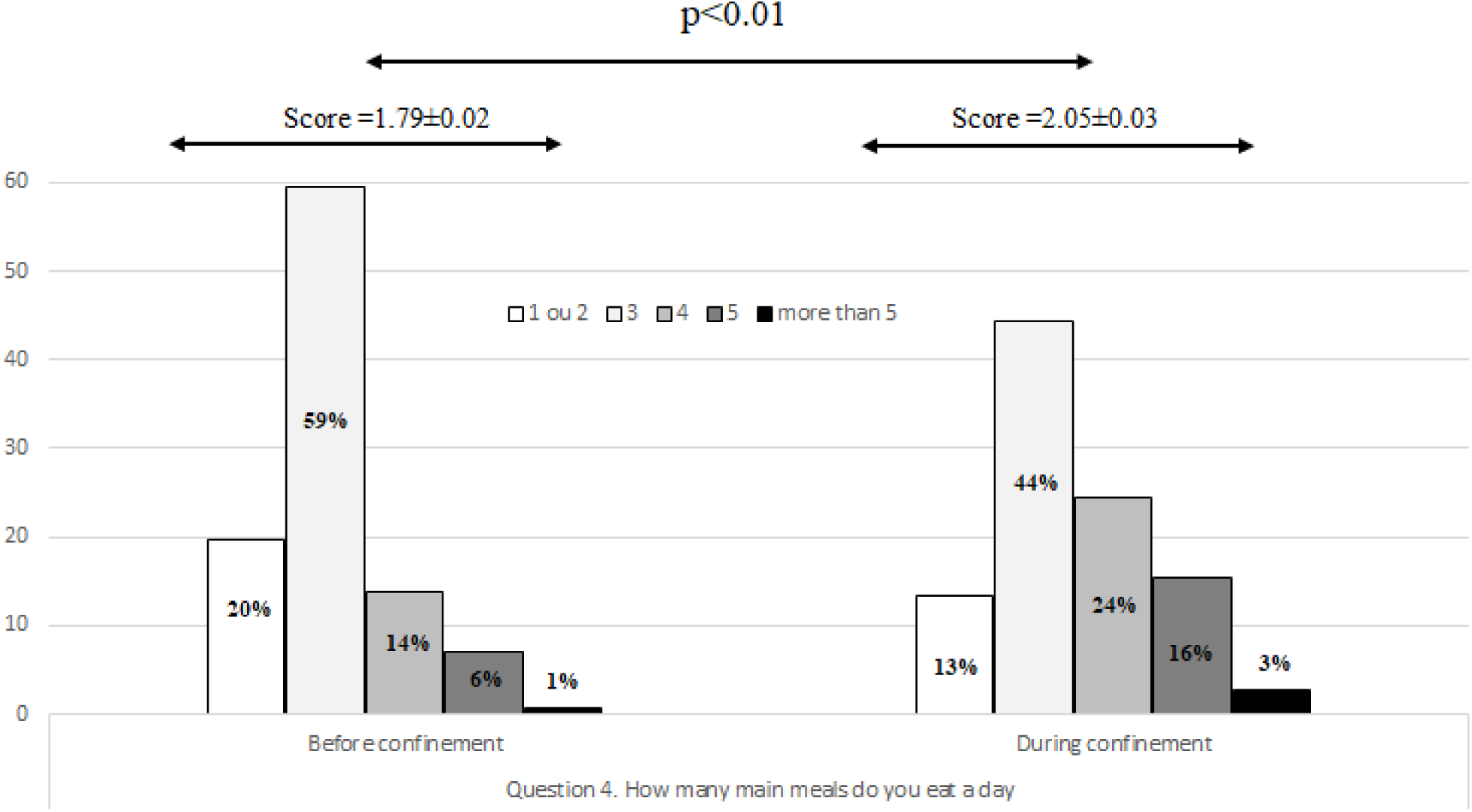
Response distribution (in %) and recorded score according to the number of main meals per day before and during confinement.

### Total score of diet

Statistical analysis reported that the total score of diet was 4.4% higher during, compared to before, home confinement (t=-10.66, p<0.001, d=0.50).

### Question 1 (Q1): Unhealthy food

The score of Q1 (consuming unhealthy food; Figure 1) was significantly higher during home confinement (t=-3.46, p<0.001, d=0.14). The percentage of responses that indicated consuming unhealthy food either most of the time or always was higher during home confinement (23.3% *vs*. 18.4% for most of the time and 10.9% *vs*. 6.2% for always).

### Question 2 (Q2): Eating out of control

The score of Q2 (eating out of control; Figure 1) was significantly higher during home confinement (t=-9.44, p<0.001, d=0.22). The percentage of responses that indicated eating out of control either most of the time or always was higher during home confinement (20.4% *vs*. 9.7% for most of the time and 9.6% *vs*. 2.3% for always).

### Question 3 (Q3): Snacking between meals

The number of snacks between meals or late-night snacking estimated by the Q3 (Figure 1) increased significantly during home confinement (t=-6.89, p<0.001, d=0.30). The percentage of responses that indicated having a snack between meals or late-night snack either most of the time or always was higher during home confinement (24.4% *vs*. 13.9% for most of the time and 15.4% *vs*. 6.4% for always).

### Question 4 (Q4): Alcohol binge drinking

The score of Q4 (binge alcohol drink; Figure 1) decreased significantly during home confinement (t=-12.16, p<0.001, d=0.58). In fact, the percentage of responses that indicated alcohol binge drinking either sometimes, most of the time or always was lower during home confinement (5.4% *vs*. 10.1% for sometimes, 1.2% *vs*. 1.8% for most of the time and 0.2% *vs*. 0.4% for always).

### Question 5 (Q5): Number of main meals/day

The number of main meals estimated by the Q5 (Figure 2) was significantly higher during home confinement (t=-5.83, p<0.001, d=0.22). The percentage of responses that indicated 4, 5 and more than 5 main meals were higher during home confinement (14.5% *vs*. 6.6% for 4 main meals, 6.3% *vs*. 2.4% for 5 main meals and 2.8% *vs*. 0.8% for more than 5 main meals)

## Discussion

This report presents the preliminary data from an online survey collected in eight languages, comparing physical activity (PA) and dietary behaviours before and during home confinement enforced as a result of COVID-19. There were 1047 replies (54% women) from Western Asia (36%), North Africa (40%), Europe (21%) and other countries (3%) which revealed that the COVID-19 home confinement has had a negative effect on all levels of PA (vigorous, moderate, walking and overall) and an increase in daily sitting time by more than 28%. Additionally, a more unhealthy pattern of food consumption (the type of food, eating out of control, snacks between meals and number of meals) was exhibited. Only alcohol binge drinking decreased significantly.

Despite the recommendation that home confinement should not hinder people from being physically active,^19^ our results show that there is a decline in all PA levels during the home confinement enforced during the COVID-19 pandemic. While the effect size is small to medium for most parameters, the 35% reduction in numbers of days per week walking is medium to large. In fact, 2.45 days is a serious change, independent from the number of walking days before confinement. However, the most prominent change was in sitting behaviour, which increased more than a full standard deviation (very large effect size: d=1.13), most likely due to the increased time that people were required to stay within the home in the quarantine. Indeed, 29% of the sample report to sit for 6-8 hours a day during confinement (vs. 24% before), a threshold area for which Patterson et al.^20^ assign increased disease and mortality risks. Far more serious was the proportion of individuals who sat for more than 8 hours a day, which increased from 16% before to 40% during confinement. Preliminary data indicate that 41% of the sample increased their sitting behaviour by only 1 hour or less, but this increased by five hours and more for 27% of the sample.

The results of this survey concur with recent studies demonstrating that the current COVID-19 home confinement could dramatically impact lifestyle activities globally, including participation in sports and PA engagement.^21^ The restrictions have reduced overall PA (number of days and number of hours) and access to exercise. In spite of an increased offering of PA guidance and classes available on social media, our results indicate that it has not been possible for individuals to adequately maintain their normal PA patterns with home activities. The decline in PA was accompanied by increased sedentary (sitting) behavior. However, the extent to which PA participation is impacted by the current COVID-19 pandemic will be linked to the stringency of the local government confinement policy. It is already been shown in China that different regional policies and socio-economic factors were associated with differences in PA.^1^ These factors need to be considered when design and promoting PA interventions for the COVID-19 pandemic.

The results of this survey also found that, in contrast to the guidance, people changed their eating behaviors, with increased consumption of unhealthy food, eating out of control, more snacking between meals and an overall higher number of main meals.^22^ Regarding dietary behaviours, there seems to be no single behavioural problem. While a medium effect size (d=0.5) was recorded for the total score, a small effect sizes was registered for the type of food, number of meals and easting out of control. The effect size for snacks between meals was medium. Binge drinking alcohol showed the largest effect size (d=0.58), but, fortunately into the healthy direction, possibly because of social isolation. In particular, younger individuals are less likely to be surrounded by other drinking peers.^23^

The negative changes in the majority of eating behaviours could be attributed to eating out of anxiety or boredom,^9^ a dip in motivation to participate in PA or maintain healthy eating^24^ or an increase in anxiety or mood driven eating.^9^ Alternative support for motivation during home confinement may be sourced from assistive technologies such as apps, streaming services and social media. To counteract poor dietary behaviours, meal planning and opting for healthy alternatives is the best approach to combating unhealthy eating habits while in confinement.^9^

The results reported in this report should be utilised for further research and development in public health promotion during the COVID-19 pandemic. Motivating people to stand up can be a first step of health promotion against sedentary behaviour and its risks. In this regard, experiences from occupational “white collar” health may be translated into the isolation situation at home. Further research should address (1) insight into sub-populations, for the development of bespoke interventions to address their needs; (2) interference of diet and PA behaviours, for improving interventions, and (3) identification of conditions for successfully maintaining a healthy lifestyle before as well as during isolation.

Although many ideas and recommendations already exist,^21,25,26^ individuals seem to need more support to effectively use the services offered and to understand the consequences of inaction. Technology and social media allow for innovative health behaviour support via fitness applications and video streaming, motivation and gamification support, and also adding the beneficial social aspect which is very important to encourage maintenance of physical activity behaviour.

## Strengths and limitations

The strengths of this research project include a survey provided in multiple languages, and which has been widely distributed in several continents. Scientists from different disciplines and many countries cooperated to make this possible. On the other hand, it should be acknowledged that there were also limitations of the low-threshold strategy in that it did not allow for narrow target groups with defined inclusion and exclusion criteria. Thus, a collection from a representative sample cannot be expected. Only during post-hoc studies, can criteria-based sub-samples can be analysed. The validity of answers is a general problem of online surveys, but we tried to face this by the differential approach described in the methods section.

## Conclusion

The preliminary results of the survey indicate a negative effect of home confinement on PA and a significant increase in sitting time with a large effect size which is indicative of a more sedentary lifestyle. These observations have potential important implications that could help PA and nutritional recommendations to maintain health during the COVID-19 pandemic.

## Data Availability

The data that support the findings of this study are available from the corresponding author, upon reasonable request.

## Acknowledgement

We thank our consortium’s colleagues who provided insight and expertise that greatly assisted the research. We thank all colleagues and peoples who believed on this initiative and helped to distribute the anonymous survey worldwide. We are also immensely grateful to all participants who #StayHome & #BoostResearch by voluntarily taken the #ECLB-COVID19 survey.

